# Tracking white and grey matter degeneration along the spinal cord axis in degenerative cervical myelopathy

**DOI:** 10.1101/2021.04.18.21255238

**Authors:** Kevin Vallotton, Gergely David, Markus Hupp, Nikolai Pfender, Julien Cohen-Adad, Michael Fehlings, Rebecca S. Samson, Claudia A. M. Gandini Wheeler-Kingshott, Armin Curt, Patrick Freund, Maryam Seif

**Affiliations:** Spinal Cord Injury Center Balgrist, University of Zurich, Zurich, Switzerland; NeuroPoly Lab, Institute of Biomedical Engineering, Polytechnique Montreal, Montreal, QC, Canada; Functional Neuroimaging Unit, CRIUGM, University of Montreal, Montreal, QC, Canada; Mila - Quebec AI Institute, Montreal, QC, Canada; University of Toronto Spine Program and Toronto Western Hospital, Toronto, Ontario, Canada; NMR Research Unit, Queen Square MS Centre, UCL Queen Square Institute of Neurology, Faculty of Brain Sciences, London, United Kingdom; Department of Brain and Behavioural Sciences, University of Pavia, Pavia, Italy; Brain Connectivity Centre, IRCCS Mondino Foundation, Pavia, Italy; Department of Neurophysics, Max Planck Institute for Human Cognitive and Brain Sciences, Leipzig, Germany; Department of Brain Repair and Rehabilitation, UCL Institute of Neurology, London, United Kingdom; Wellcome Trust Centre for Neuroimaging, UCL Institute of Neurology, London, United Kingdom

## Abstract

**Objective:** To determine tissue-specific neurodegeneration across the spinal cord in patients with mild-moderate degenerative cervical myelopathy (DCM).

**Methods:** Twenty-four mild-moderate DCM and 24 healthy subjects were recruited. In patients, a T2-weighted scan was acquired at the compression site, while in all participants a T2*-weighted and diffusion-weighted scan was acquired at the cervical level (C2-C3) and in the lumbar enlargement (i.e. rostral and caudal to the site of compression). We quantified intramedullary signal changes, maximal canal and cord compression, white (WM) and grey matter (GM) atrophy, and microstructural indices from diffusion-weighted scans. All patients underwent clinical (modified Japanese Orthopaedic Association (mJOA)) and electrophysiological assessments. Regression analysis assessed associations between MRI readouts and electrophysiological and clinical outcomes.

**Results:** Twenty patients were classified with mild and four with moderate DCM using the mJOA scale. The most frequent site of compression was at C5-C6 level with maximum cord compression of 4.68±0.83 mm. Ten patients showed imaging evidence of cervical myelopathy. In the cervical cord, WM and GM atrophy and WM microstructural changes were evident, while in the lumbar cord only WM showed atrophy and microstructural changes. Remote cervical cord WM microstructural changes were pronounced in patients with radiological myelopathy and associated with impaired electrophysiology. Lumbar cord WM atrophy was associated with lower limb sensory impairments.

**Conclusion:** Tissue-specific neurodegeneration revealed by quantitative MRI, already apparent across the spinal cord in mild-moderate DCM prior to the onset of severe clinical impairments. WM microstructural changes are particularly sensitive to remote pathologically and clinically eloquent changes in DCM.

## Introduction

Degenerative cervical myelopathy (DCM) is the most common cause of non-traumatic spinal cord injury and can lead to spinal cord dysfunction. Degenerative changes of the spine lead to a progressive stenosis of the cervical spinal canal[1,2] with ensuing spinal cord compression. Cord compression triggers a cascade of pathophysiological processes (ischemia, inflammation, neuronal and oligodendroglial apoptosis) at the site of compression producing irreversible neural tissue damage (cervical myelopathy)[3] as well as secondary anterograde and retrograde degeneration of spinal pathways[4] above and below the compression site.[5–7]

To detect tissue at risk and prevent irreversible tissue damage in the early stages of DCM, there is a pressing need to determine tissue integrity and the underlying pathophysiology[3] at and remote to the site of spinal cord compression. Quantitative MRI (qMRI) techniques can provide biomarkers sensitive to spinal cord tissue integrity[8] and underlying pathology in DCM[7,9] and help clinical decision making (i.e. early surgery vs. conservative treatment).

This study aims therefore to track compression-induced neurodegeneration across the spinal cord axis using an advanced qMRI protocol[10,11] that includes tissue-specific volumetric and microstructural indices derived from structural and diffusion-weighted imaging. We hypothesized that (i) tissue-specific neurodegeneration extends from the site of compression in the rostral and caudal directions in mild-moderate DCM, (ii) remote white matter microstructural changes are evident prior to irreversible tissue damage at the site of compression (i.e. radiological evidence of cervical myelopathy), and (iii) spinal cord pathology is clinically eloquent remote to the site of cord compression.

## Methods

### Standard protocol approvals, registrations, and patient consents

The study protocol was performed in accordance with the Declaration of Helsinki, approved by the local Ethics Committee of canton Zurich (EK-2010-0271). Informed written consent was obtained from each participant before study enrolment.

### Participants

Twenty-four mild (n=20, mJOA) ≥15) to moderate (n=4,12 ≤mJOA≤14) DCM patients (AIS C-E, mean (±SD) age = 54.92±11.00 years, 8 female) and 24 healthy controls (mean age (±SD) = 41.6±15.4 years, 4 female) were recruited from the Spinal Cord Injury Centre outpatient clinic. Inclusion criteria were a spinal canal stenosis observed in MRI and the clinical diagnosis of DCM[2] no previous spine operations, no head or brain lesions, no pre-existing neurological and mental disorder, no MRI contraindications and 18 < age < 70 years.

### Clinical assessments

All patients underwent a clinical examination specific to DCM including the modified Japanese Orthopaedic Association (mJOA) score[12] and the Nurick scale.[13] The mJOA score is a validated disease-specific outcome measurement quantifying clinical impairment in the upper and lower limbs as well as sphincter function on an 18-point scale as follows,[12] where a score of 18 reflects no impairment and lower scores indicate a progressively greater degree of disability and functional impairment. The Nurick classification[13] was conducted to grade patients into five different categories, from 0 to 5, where a grade of 0 indicates no evidence of spinal cord involvement to the patient’s symptoms and a grade of 5 indicates that the patient is chair-bound or bedridden. In addition, the International Standards for Neurological Classification of Spinal Cord Injury (ISNCSCI) protocol[14] was performed, which discerns detailed assessments of the upper and lower extremity motor function, as well as light-touch and pinprick sensation across all spinal segments.

### Electrophysiological measurements

The cervical dermatomal somatosensory evoked potentials (SSEPs) measurements were conducted in patients at the C6 and C8 dermatomes according to the standard protocol of the European Multi-centre Study About Spinal Cord Injury (EMSCI).[15]

### Image acquisition

Participants were positioned head-first supine on a 3T MRI system (Skyra^Fit^ Siemens Healthcare, Erlangen, Germany). Radio Frequency (RF) excitation was performed using the body coil and detection was achieved using a combination of a twelve-channel head-coil, four-channel neck-coil, and 24-channel spine matrix. Additionally, an MRI protocol was performed in patients to assess the extent of the lesion, consisting of an anatomical 2D sagittal scan and an axial T2-weighted scan based on a turbo spin echo sequence. The MRI parameters of the sagittal T2-weighted scan were as follows: repetition time (TR)=3500 ms, echo time (TE)=84 ms, flip angle=160°, field of view (FOV)=220×220 mm^2^, and in-plane resolution=0.34×0.34 mm^2^ readout bandwidth =260 Hz/pixel, acquisition time=1.37 min; the MRI parameters for the axial T2-weighted scan were as follows: TR=5510 ms, TE=96 ms, flip angle=150° FOV=160×160 mm^2^, in-plane resolution=0.6×0.6 mm^2^, readout bandwidth=283 Hz/pixel, acquisition time=1.57 min.

To quantify atrophy at the cervical cord (C2-C3) and at lumbar enlargement (T11-L1) levels, a T2*-weighted three-dimensional (3D) sequence (multiple echo data image combination; MEDIC) was performed. The 10 axial–oblique slices were centred at the C2 intervertebral disc on the cervical level and on the widest point of the lumbar cord enlargement as appearing in a localizing sagittal T2-weighted image.[16] The T2*-weighted images resulted in four axial 3D volumes of the cervical and lumbar cord with in-plane resolution of 0.5×0.5 mm^2^, slice-thickness=5 mm and FOV=162 ×192 mm^2^. TR=44 msec for the cervical cord and 55 msec for the lumbar cord imaging, TE=19 msec, flip angle = 1°, and readout bandwidth = 260 Hz/pixel acquisition time= 3.30 min at the cervical and 4.22 min at the lumbar cord level. These sequences are based on the spine generic consensus protocol.[10,11]

To quantify microstructural changes of the spinal cord at the cervical and lumbar level, a diffusion-weighted imaging (DWI) sequence was performed based on the FOV single-shot spin-echo echo planar imaging (EPI) method with cardiac-gating (based on finger pulse oximetry) resulting in 60 diffusion-weighted images (b-value=500 sec/mm^2^) and six b_0_-weighted images. DWI MR parameters were as follows: TR=350 msec, TE=73 msec, slice thickness=5 mm with 10% inter-slice gap, bandwidth=812 Hz/pixel, FOV=133 × 30 mm^2^, in-plane resolution=0.8×0.8 mm^2^, and nominal total acquisition time of 6.2 min.

As a result of motion artefacts at cervical level, 3 patients and one healthy control were excluded from atrophy analysis, and 3 patients and 5 healthy controls from diffusion analysis. At the lumbar cord level, 5 patients and 3 healthy controls were excluded from atrophy assessment and 5 patients and 5 healthy controls were excluded from diffusion assessment.

### Image processing

#### Processing of T2-weighted MRI at the compression site

T2-weighted MRI was used to determine the exact level(s) of spinal canal stenosis and intramedullary signal hyperintensity as sign of myelopathy (radiological evidence of cervical myelopathy) along the cervical spinal cord. Next, the mid-sagittal slice of the T2-weighted images was assessed to determine the maximum spinal cord compression (MSCC), the maximum canal compromise (MCC) and to calculate signal intensity of the signal hyperintensity on T2-weighted images.[17]

A region of interest (ROI) with area of 0.05 cm^2^ was placed on the spinal cord at the compression site, as well as above and below the compression site as reference areas. Next, the signal intensity ratio was calculated against the average of the reference ROIs.[18]

To show the distribution frequency of lesions in the patient group voxel-wise, hyperintense voxels were manually segmented on axial slices by an experienced rater (KV) using FSLeyes from FMRIB software library v6.0. Next, on the same sagittal T2-weighted images the total spinal cord area was segmented applying the Spinal Cord Toolbox (SCT) (version 3.2.2).[19] The lesion and spinal cord masks and T2-weighted images were normalized to the T2-weighted PAM50 spinal cord template[20] using a slice-wise non-linear registration using SCT. To illustrate the distribution of lesions over vertebral levels and in the axial plane, a group-level lesion frequency map was created by applying the forward-transformation (native space to PAM50, nearest-neighbour interpolation) on the individual lesion masks, followed by averaging across subjects on a voxel-wise basis. In the resulting lesion frequency map, the voxel intensity represents the frequency of a lesion (%) occurring in the corresponding voxel.

#### Processing of T2*-weighted MRI at cervical and lumbar cord level

The serial longitudinal registration algorithm available in SPM12[21] was applied to all T2*-weighted to average the images accounting for intra-participant motion. Jim 7.0 software was used to semi-automatically segment the cross-sectional cord (SCA), grey (GMA) and white matter (WMA) area using an active-surface model after setting a marker in the centre of the cord in each of the 10 contiguous slices. GMA was manually segmented. Next, WMA was obtained by subtracting GM masks from the total SC masks. Of note, at the lumbar enlargement level, three slices with the largest SC cross-sectional area was selected[16] to ensure the anatomical level while at the cervical cord all slices were considered for the further analysis.

#### Processing of diffusion weighted MRI at cervical and lumbar cord level

Processing of DWI data was carried out with a modified version of the MATLAB-based Artefact Correction in Diffusion MRI (ACID) toolbox[22] within SPM12 optimized for the spinal cord. First, we reduced the in-plane FOV to 30×30 mm^2^ to include only spinal cord tissue. Eddy Current Motion Correction (ECMOCO) algorithm of ACID toolbox was applied[23] to correct for intra-participant motion and eddy-current artefacts. The diffusion tensor model was fitted to the DWI data by applying a robust tensor fitting algorithm in ACID toolbox.[22,24] The DWI fitting resulted into Diffusion Tensor Imaging (DTI) maps including fractional anisotropy (FA), mean diffusivity (MD), axial diffusivity (AD) and radial diffusivity (RD). All DTI maps were registered to the corresponding T2*-weighted image applying a nonlinear transformation (BSplineSyn algorithm)[25] implemented with regularization across slices and the b_0_ image as a source image to drive the registration. Mean FA, AD, MD, and RD were extracted from the SC, GM, and WM binary masks, using the ACID toolbox.

### Statistical analysis

Statistical analysis was performed using Stata 16 (Stata-Corp LP, College Station, TX). The mean age was higher in DCM patients compared to healthy controls (mean±SD), DCM: 54.9±11.0, healthy control: 41.6±15) (t-test, p=0.001), thus age was considered as covariate of no interest in our linear regression models. Macrostructural morphometric measurements (i.e. SCA, GMA and WMA) were compared between healthy controls and DCM patients by means of one-tailed t-test. Next, microstructural DTI indexes (FA, AD, MD and RD) within cervical and lumbar cord were compared between groups using two-sample t-test (unpaired, one-tailed, p<0.05).

Patients were divided into two groups depending on the presence versus absence of T2-hyperintensity signal in the cervical cord as a sign of cervical myelopathy. Measurements of MSCC and MCC, volumetric and microstructural MRI readouts as well as clinical scores were compared between groups using two-sample t-test (unpaired, one-tailed, p<0.05). Finally, linear regression analysis in Stata was performed to reveal possible relationships between volumetric and microstructural MRI readouts in cervical and lumbar cord and clinical and electrophysiological outcomes, adjusted for age, using a level of significance set to p<0.05.

## Results

### Demographic, clinical, electrophysiological, and radiologic characteristics

Twenty-four mild-moderate DCM patients and 24 healthy controls were recruited and underwent MRI protocol. Out of the 24 DCM patients, 20 patients had mild (mJOA≥15) and 4 had moderate DCM (12≤mJOA≤14). Moreover, 8 patients had a Nurick grade of 0/5, 13 patients had a Nurick grade of 1/5 and 3 patients had a Nurick grade of 2/5. Within the ISNCSCI examination, the light-touch score was (mean±SD) 28.56±4.79 pts for the upper and 30.67±2.76 pts for the lower extremities (max. value 32 points). The pinprick score of the upper extremities (max. value 32 points) was 27.8±4.8 pts and the pinprick score of the lower extremities was 30.67±2.76 pts. The upper and lower extremity motor scores (max. value 50 points) were 48.04±6.5 pts, 48.5±7.0 pts, respectively.

Cervical dermatomal SEPs showed values considered in the normal range in clinical practice:[26,27] a sensory threshold of 3.20±0.64mA for C6 and 3.28±0.45 mA for C8, a pain threshold of 18.81±6.71 mA for C6 and 16.37±5.39 mA for C8, an amplitude of 1.39±0.81 mV for C6 and 1.17±0.61 mV for C8, a N1 latency of 24.21±1.62 msec for C6 and 24.89±1.89 msec for C8 and a P1 latency of 23.30±1.99 msec for C6 and 29.15±6.5 msec for C8.

The maximal cervical spinal stenosis was mostly observed at cervical level C5-C6 (n=17) (Table 1). Of the 24 patients, 16 had multi-segmental cervical spinal canal stenosis. Signal hyperintensity of the cord on the T2-weighted images was observed in ten patients (Fig. 1A & B). The frequency map of T2-hyperintensity signal (“myelopathy”) across the cervical cord in DCM patients with myelopathy has been shown for 10 patients in figure 1C. The map shows that the T2-weighted hyperintensity is more frequent on the C4-C6 levels in these patients (Fig. 1C).

**Table 1.**
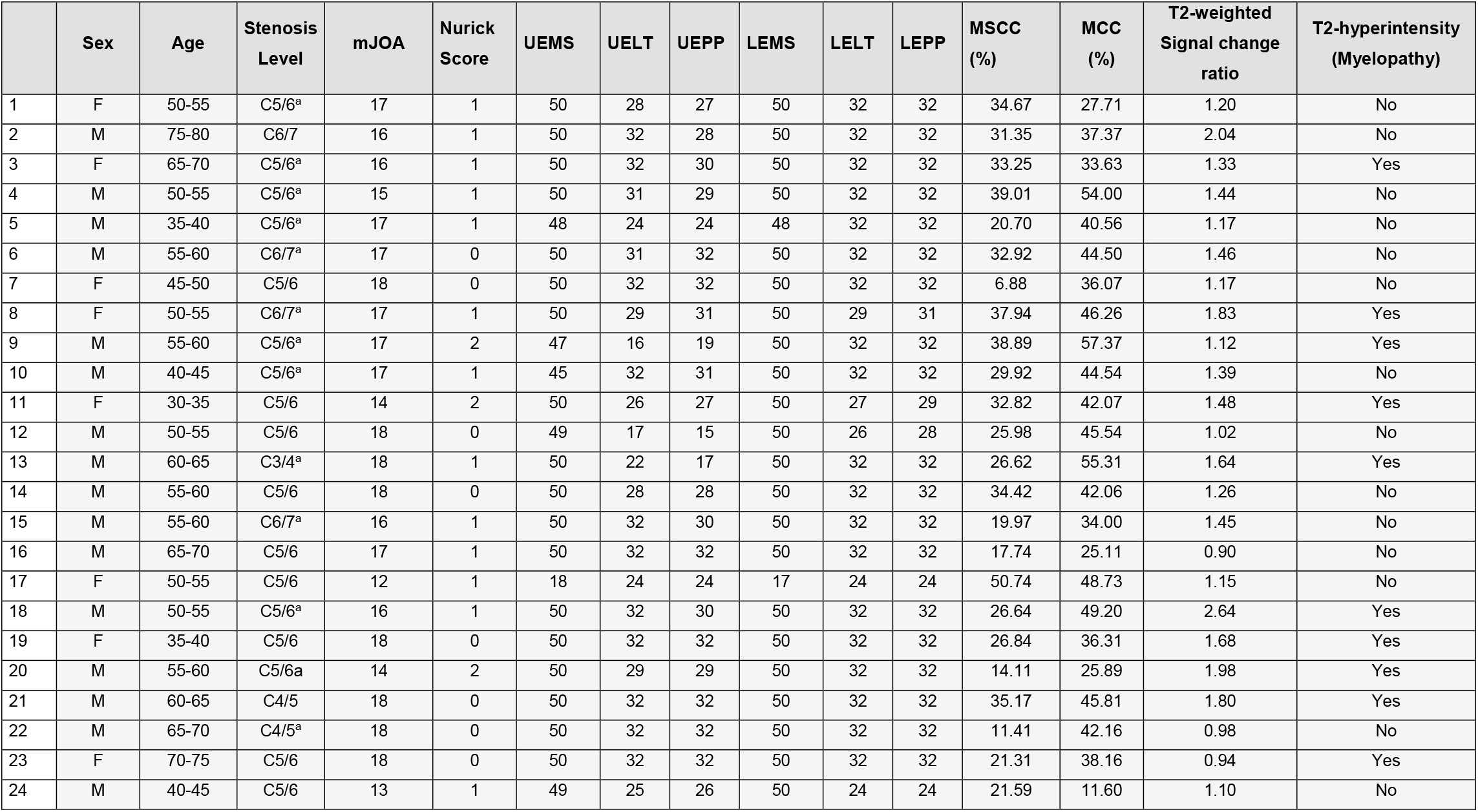
Clinical data of patients with degenerative cervical myelopathy (DCM). mJOA = modified Japanese Orthopaedic Association [max. 18 points]. UEMS = upper extremity motor score [max. 50 points]. UELT = upper extremity light-touch [max. 32 points]. UEPP = upper extremity pinprick [max. 32 points]. LEMS = lower extremity motor score [max. 50 points]. LELT = lower extremity light-touch [max. 32 points]. LEPP = lower extremity pinprick [max. 32 points]. MSCC = maximum spinal cord compression. MCC = maximum canal compromise. ^a^ multi-segmental cervical spine stenosis. Age has been reported in a range (5-year range minimum).

**Figure 1:**
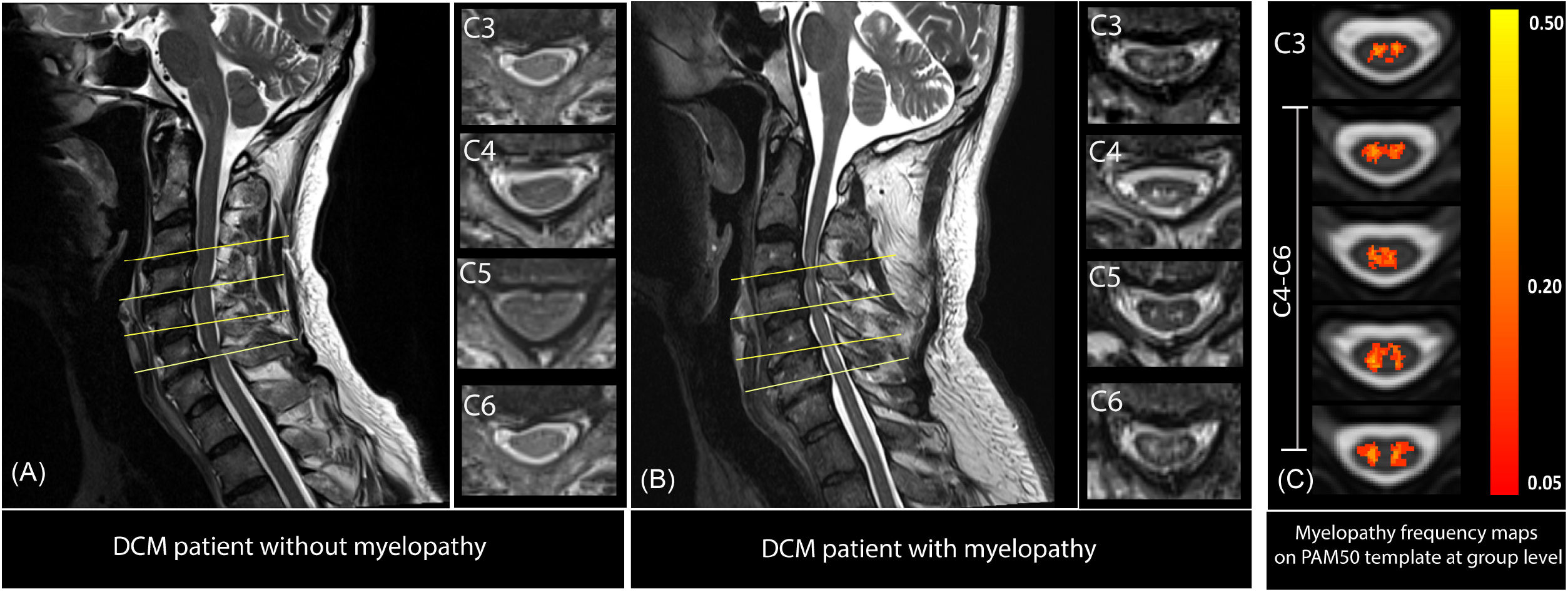
Overview of the site of compression in DCM patients (A) without and (B) with radiological evidence of cervical myelopathy. (C): Frequency map of T2-hyperintensity signal (“cervical myelopathy”) across the cervical cord in DCM patients with myelopathy. Colour code: red= low frequency of hyperintense signal, yellow=high probability of hyperintense signal. The map was composed based on all DCM patients showing T2-hyperinsity signal (n=10) and overplayed on PAM50 template[20].

DCM patients with radiological evidence of cervical myelopathy had reduced MSCC at their maximal compression site in comparison with patients without cervical myelopathy (DCM with radiological evidence of myelopathy n=10: MSCC=4.10±0.24 mm, DCM without radiological evidence of myelopathy n=14: MSCC=4.93±0.27 mm (p=0.02)) and reduced MCC (DCM with myelopathy MCC=6.29±0.31mm, DCM without myelopathy MCC=7.03±0.25 mm, p=0.04). There were no significant differences between clinical scores (e.g. mJOA, Nurick, and INSCSCI scores) in patients with and without radiological evidence of cervical myelopathy.

### Spinal cord neurodegenerations at cervical and lumbar cord

We first confirmed findings reported in previous studies[7,28] that at cervical cord level (C2-C3) the SCA was decreased (Δ=-12.0%, p<0.001) in DCM patients compared with healthy controls (Table 2). Sub-segmentation of the spinal cord showed that both the GMA and WMA were decreased in DCM patients in comparison to healthy controls (GMA: Δ=-10.4%, p<0.001, WMA: Δ=-12.1%, p<0.001).

**Table 2.** Tissue-specific cross-sectional areas and diffusion tensor imaging scalar values in the cervical cord at level C2-C3 and in the lumbar enlargement in patients with degenerative cervical myelopathy and healthy controls. SCA: spinal cord area, GMA: grey matter area, WMA: white matter area. ROI: region of interest, FA: fractional anisotropy, MD: mean diffusivity, AD, axial diffusivity, RD: radial diffusivity.

| Cervical cord at C2-C3 level |  |  |  |  |  | Lumbar cord enlargement |  |  |  |
| --- | --- | --- | --- | --- | --- | --- | --- | --- | --- |
| MRI readout | ROI | Controls | Patients | Difference | *p* Value | Controls | Patients | Difference | *p* Value |
| Cross-sectional area, mm<sup>2</sup> | SCA | 87.4±8.3 | 76.9±6.3 | -12.0% | 0.0002 | 64.2±7.0 | 58.7±5.7 | -8.60% | 0.014 |
|  | GMA | 12.6±1.2 | 11.3±0.9 | -10.4% | 0.0007 | 18.7±2.7 | 18.3±3.0 | -2.0% | 0.69 |
|  | WMA | 74.8±7.4 | 65.8±6.0 | -12.1% | 0.0003 | 45.5±6.4 | 40.4±4.7 | -11.30% | 0.01 |
| FA | WM | 0.70±0.05 | 0.67±0.04 | -4.6% | 0.01 | 0.55±0.03 | 0.53±0.04 | -2.90% | 0.08 |
|  | GM | 0.62±0.05 | 0.61±0.05 | -2.4% | 0.15 | 0.36±0.05 | 0.36±0.07 | 1.1% | 0.42 |
| MD, µm<sup>2</sup>/ms | WM | 1.07±0.10 | 1.19±0.12 | 10.58% | 0.001 | 1.14±0.09 | 1.26±0.18 | 9.5% | 0.01 |
|  | GM | 1.04±0.17 | 1.1±0.18 | 4.17% | 0.21 | 0.92 ±0.05 | 0.91±0.06 | 0.43% | 0.83 |
| AD, µm<sup>2</sup>/ms | WM | 2.10±0.24 | 2.24±0.23 | 6.8% | 0.01 | 1.91±0.16 | 2.06±0.26 | 7.1% | 0.04 |
|  | GM | 1.86±0.22 | 1.91±0.36 | 2.62% | 0.27 | 1.34±0.11 | 1.33±0.17 | 0.45% | 0.45 |
| RD, µm<sup>2</sup>/ms | WM | 0.55±0.09 | 0.69±0.17 | 14.50% | 0.001 | 0.76±0.09 | 0.86±0.14 | 11.0% | 0.02 |
|  | GM | 0.63±0.16 | 0.67±0.12 | 4.86% | 0.23 | 0.73±0.05 | 0.73±0.06 | 0.8% | 0.72 |

Considering microstructural indices, FA was decreased in the cervical WM in DCM patients compared to healthy controls (Δ=-4.6%, p=0.01). In the WM of the cervical cord, MD, AD and RD was increased in DCM patients compared to healthy controls (MD: Δ= +10.58%, p=0.001, AD: Δ=+6.8%, p=0.01, RD: Δ=+14.50%, p=0.001;) (Table 2).

At the lumbar level, SCA (Δ=-8.6%, p=0.014) and WMA (Δ=-11.30%, p=0.01) were decreased in DCM patients compared with healthy controls significantly, whereas GMA was similar in both groups (p=0.69) (Table 2, Fig. 2). Microstructural indices showed that MD, AD, and RD were significantly increased in DCM patients compared with healthy controls in WM (MD: Δ=9.5% p=0.01; AD: Δ=7.1%, p=0.04. RD: Δ=11%, p=0.02). However, decrease of FA in WM of DCM patients compared to healthy controls was not significant (p=0.42) and DTI indices did not change in the GM (Table 2 and Fig. 2).

**Figure 2:**
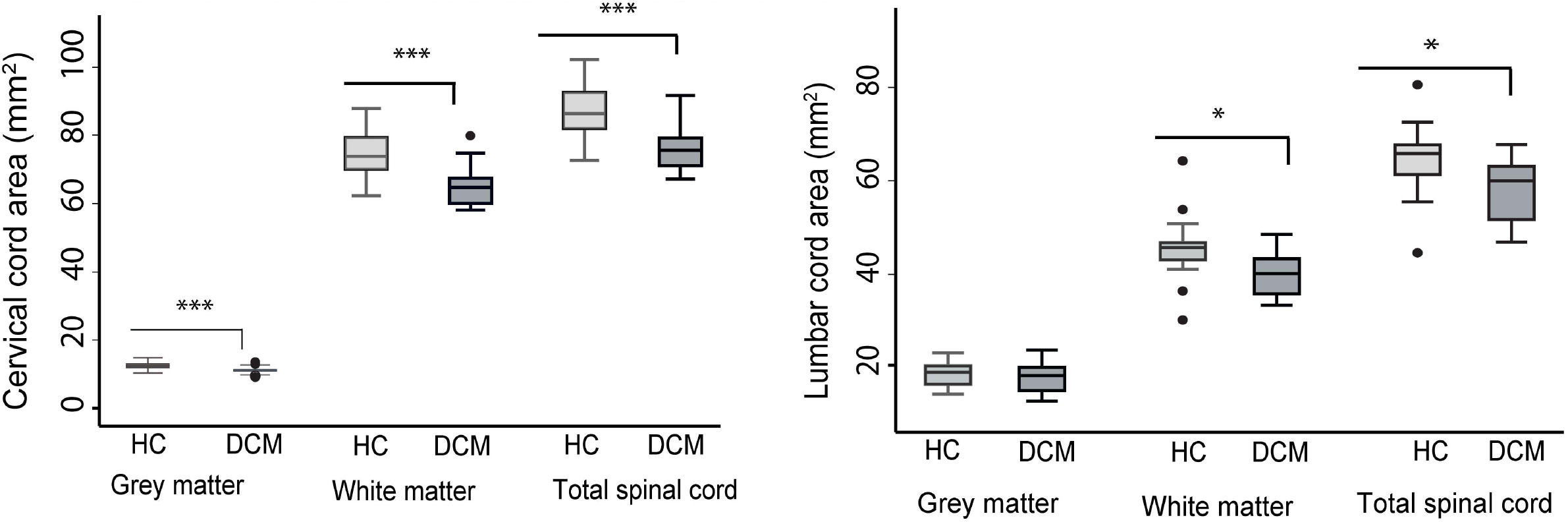
Box plots of cross-sectional areas of total spinal cord, grey and white matter areas in the cervical and lumbar cord of both DCM patients and healthy controls. \*\*\**p*<0.001, \**p*<0.05.

### Associations between cervical and lumbar cord neurodegeneration

In DCM patients, total cross-sectional SC and WMA at the cervical C2-C3 level predicted the corresponding volumetric changes at the lumbar level (p<0.001, *r*=0.78; p=0.002, *r*=0.70 respectively). However, no association was found between GMA at cervical level (C2-C3) and in the lumbar cord enlargement.

In healthy controls, volumetric measurements at C2-C3 level were not associated with the corresponding measurements in the lumbar cord (SCA: p=0.17, *r*=0.31; WMA: p=0.06, *r*=0.42) (Fig. 3). The DTI indices measured at C2-C3 level were not associated with the DTI indices measured in the lumbar cord in both DCM patients and healthy controls.

**Figure 3:**
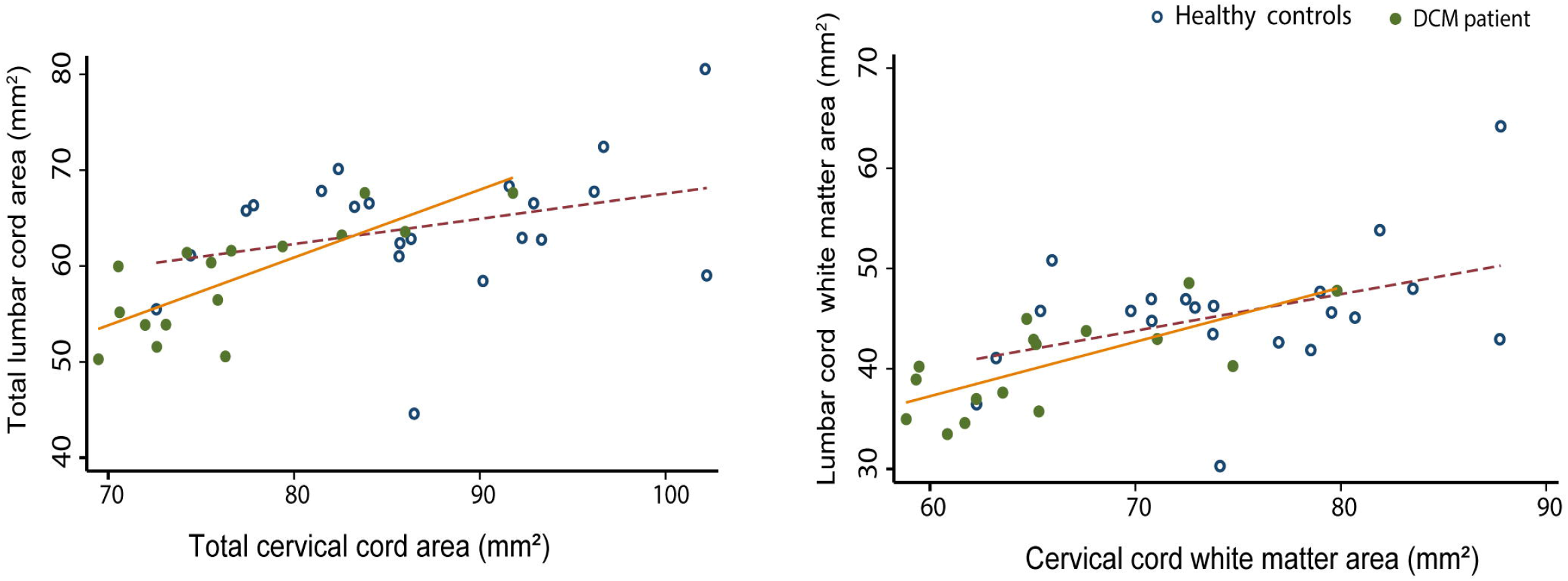
Linear regression models of spinal cord cross-sectional areas in cervical and lumbar level: the total spinal cord and white matter areas in both DCM patients and healthy controls were shown. DCM patients: cervical total spinal cord area: p<0.0001, *r*=0.78; white matter area: p=0.002, *r*=0.70). Healthy controls: cervical total spinal cord area: p=0.168, *r*= 0.31; white matter area: p=0.06, *r*=0.42

### Effects of myelopathy on remote neurodegeneration

Patients with radiological evidence of cord myelopathy (i.e. T2-weighted hyperintensity) showed reduced FA (DCM without myelopathy=0.68±0.03, DCM with myelopathy=0.63±0.05, Δ=-7.35%, p=0.005), increased RD (DCM without myelopathy =0.62±0.07, DCM with myelopathy=0.70±0.12, Δ=+22.06%, p=0.03) and increased MD (DCM without myelopathy MD= 1.17±0.13; DCM with myelopathy MD=1.22±0.11, Δ=4.27%, p>0.05) (Fig. 4) in cervical WM compared to patients without myelopathy. At the lumbar cord level, no significant differences in DTI indices were found between patients with or without radiological evidence of cord myelopathy.

**Figure 4:**
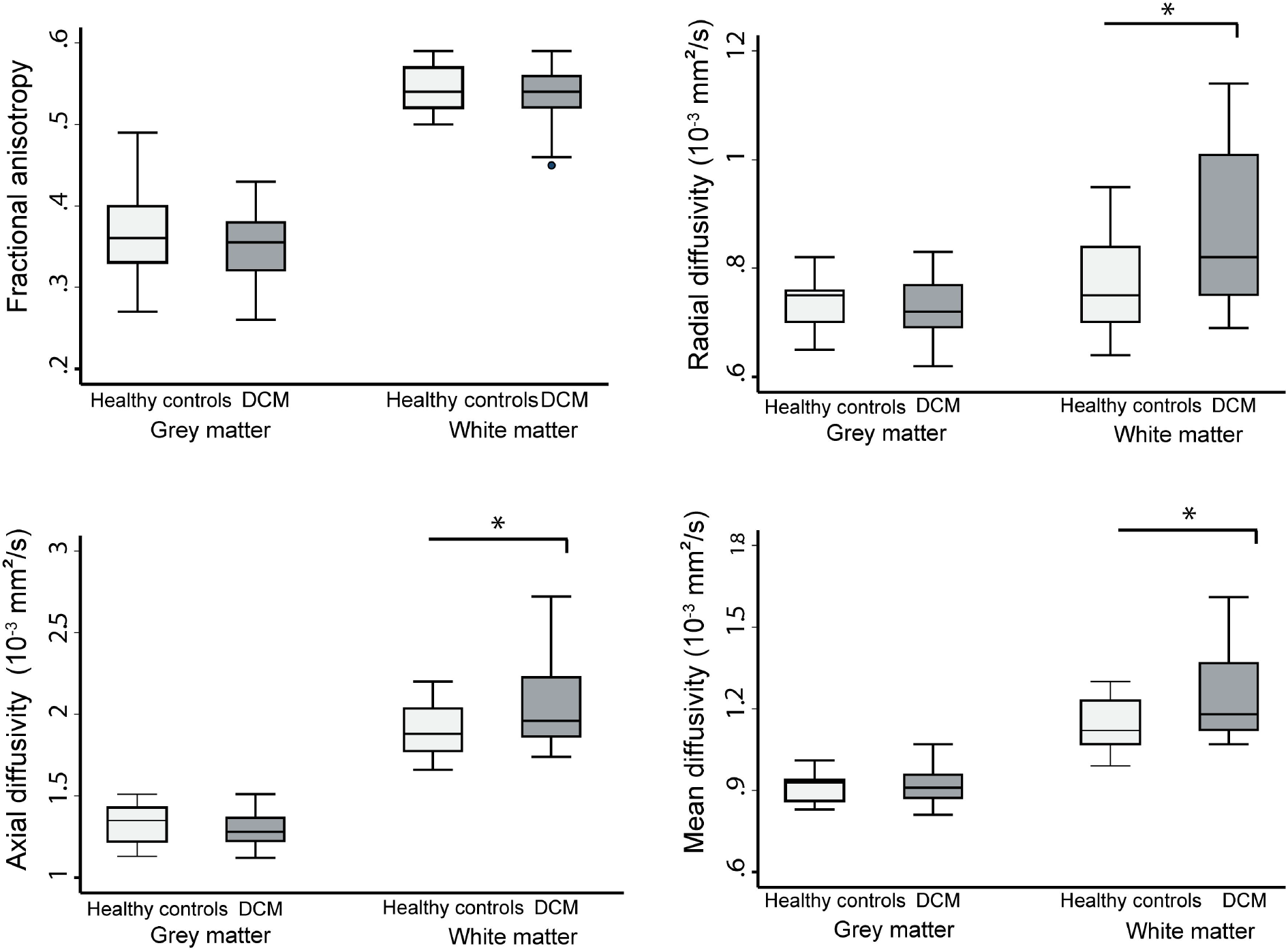
Box plots of lumbar cord diffusion tensor imaging metrics of grey and white matter of both DCM patients and healthy controls. \**p*<0.05.

**Figure 5:**
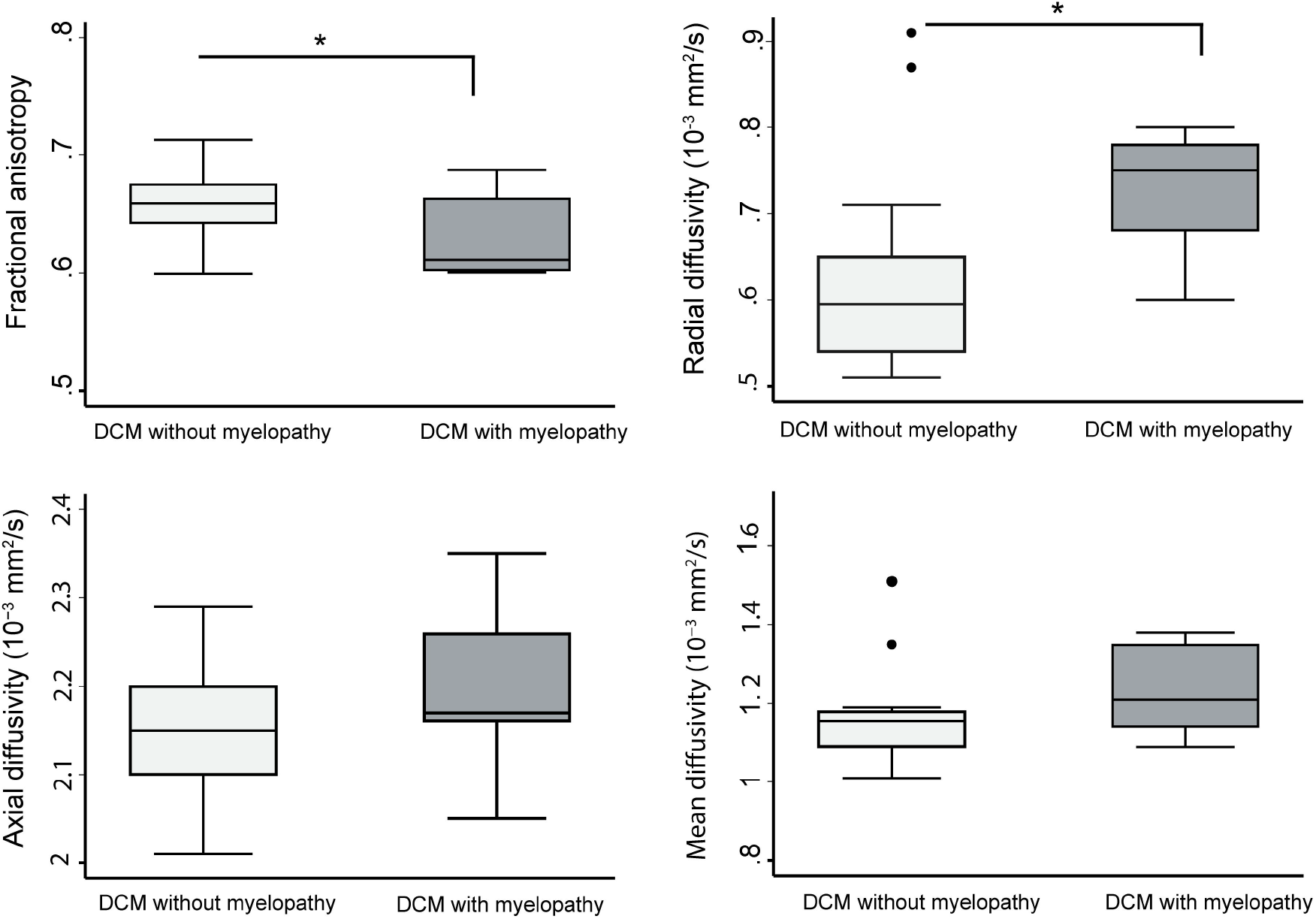
Box plots of diffusion tensor imaging (DTI) metrics of white matter in cervical (C2-C3) level of DCM patients with and without radiological signs of myelopathy.

### Associations between MRI readouts and clinical outcome

Increased MD in the cervical cord was associated with lower C6 and C8 dSEP pain thresholds (C6: n=16, p=0.031, r=-0.56, C8: n=16, p=0.01, r=-0.62). Increased AD was associated with lower pain threshold dSEP at C8 level (n=16, p=0.007, r=-0.65). Lumbar WM atrophy was associated with light touch score (p<.001, *r*=0.92) and pinprick score (p=0.001, *r*=0.86) of the lower extremities. No significant correlations were found between lumbar DTI indices and clinical impairment.

## Discussion

This study shows compression-induced neurodegeneration along the spinal cord axis in the early stages of DCM, even prior to the onset of severe functional impairments. The magnitude of cervical cord neurodegeneration revealed by microstructural indices in WM was more pronounced in patients with radiological evidence of cervical myelopathy (i.e. T2-weighted hyperintensity). Crucially, qMRI was more sensitive to myelopathy-induced changes compared to conventional MRI and clinical measures. Thus, the qMRI protocol complements standard clinical measures for diagnostic purposes,[29] revealing microstructural changes of the spinal cord in early DCM stages.

The majority of included DCM patients showed only mild clinical impairment based on both the mJOA classification and the detailed assessments of the motor and sensory function within the ISNCSCI examination. Electrophysiological recordings (i.e. cervical dermatomal SEPs) were within normal ranges or very small changes in all patients. In contrast, conventional MRI demonstrated signs of irreversible intramedullary signal changes (i.e. T2-weighted hyperintensity, reflecting cervical myelopathy) in 42% (i.e.10/24) of the patients at the site of maximal compression. While standard clinical MRI scans did not show any sign of tissue damage (i.e. normal appearing spinal cord in T2-weighted MRI) remote from the level of cord compression, qMRI revealed degeneration in cervical cord WM[7,28] as well as damage of WM in the lumbar enlargement regardless of the extent of spinal cord compression. Pathophysiological WM changes above and below the level of compression may result from secondary induced anterograde and retrograde degeneration of spinal pathways. While measurements in the cervical cord are only some segments (one to 4 segments) apart from the site of spinal cord compression, lumbar cord measurements are at least 13 segments away from the compression site, emphasizing remote alterations of the neural network away from the site of spinal cord injury. Interestingly, GM atrophy was observed only in the cervical cord. This may be due to the mechanically-induced restriction of the microvasculature and blood supply impairments[30,31] that affects more the cord in the proximity of the site of compression[7] where anterior horn cell loss and apoptosis have been reported[32,33] while in the lumbar cord this remains unaffected.[34] Thus, pathophysiological changes occurring in the lumbar cord must relate to long distance tract degeneration from cervical compression;[4,6,35] this hypothesis is in line with human post-mortem findings, where demyelination of both descending and ascending WM tracts in the lesion epicentre of DCM patients was found.[32] In accordance with these neuropathological reports, our DTI indices revealed microstructural changes in the atrophied cervical and lumbar WM. Above and below the level of compression, RD, AD, and MD increased, supporting demyelination as an integral part of the underlying neurodegenerative process.[36] These results are in line with previous studies[4,34,37] that highlighted the potential role of demyelination in neurodegeneration of the spinal cord in DCM patients.

The magnitude of cervical cord atrophy above the compression site was correlated with atrophy occurring in the lumbar enlargement, i.e. those patients with severe atrophy above the compression site showed also severe atrophy below the compression site. This could imply a common pathophysiological process causing neurodegeneration above and below the lesion site, with the compression site being the common denominator. In healthy controls, no correlations were observed between cervical and lumbar cord volumetric parameters, supporting our results as being an epiphenomenon of the DCM pathophysiology. Since those results are independent of the compression site measurements, further characterization of patients exhibiting more remote neurodegeneration could shed light on remote and targetable pathophysiological changes in DCM.

While microstructural changes of the cervical cord (C2-C3) detected by DTI were associated with lower sensory and pain thresholds in C6 and C8 dermatomal SEPs, the mJOA scores were not associated with sensory impairments. Additionally, atrophy of the lumbar cord in DCM patients was correlated with sensory lower limb impairment. Interestingly, MRI readouts at the site of compression were not associated with motor or sensory outcomes or with cervical dermatomal SEPs. This suggests that advanced qMRI methods have an enhanced sensitivity to pathological changes and the potential to identify clinically eloquent spinal cord neurodegeneration in DCM patients with mild impairments. Hereby, qMRI can provide more sensitive tools to complement current standard clinical classification methods. These results are in line with studies before, reporting tissue changes in a multimodal MRI protocol in the cervical cord in DCM patients.[5,6,28,38]

### Limitations

DCM patients were on average 14 years older than healthy controls, which may affect the compensatory effects like aging on the microstructural changes. However, age was considered as a covariate in statistical analysis to reduce age-related effects. As all electrophysiological (i.e. dSEP) measurements were within normal limits and the patients were only mildly affected, statistical analysis (i.e. correlation of qMRI readouts to the clinical data) may be affected by a ceiling effect of the clinical data.

In conclusion, this study shows that tissue-specific neurodegeneration is already apparent above and below the site of compression in DCM patients with mild to moderate clinical symptoms based on mJOA score; its magnitude being more pronounced in patients with radiological evidence of cervical myelopathy. Therefore, qMRI readouts are sensitive to remote pathological and clinical eloquent changes across the spinal cord axis prior to onset of sever clinical impairments in DCM patients. Thus, evolving clinical impairment in DCM patients are not entirely due to focal cervical cord pathology, but implies complex degeneration that occur across the spinal cord axis.[39,40]

## Data Availability

The authors certify they have documented all data, methods, and materials used to conduct the research presented. Anonymized data not published within this article will be made available by request from any qualified investigator.

## Acknowledgements

We would like to thank all subjects participating in our study who gave generously of their time, the staff of the Department of Radiology for scanning subjects as well as the Spinal Cord Injury Center Balgrist for the support in patient recruitment.

## References

1 Nouri A, Tetreault L, Singh A, et al. Degenerative cervical myelopathy: Epidemiology, genetics, and pathogenesis. Spine (Phila. Pa. 1976). 2015; 40:E675–93. doi:10.1097/BRS.0000000000000913

2 de Oliveira Vilaça C, Orsini M, Leite MAA, et al. Cervical spondylotic myelopathy: What the neurologist should know. Neurol. Int. 2016; 8:69–73. doi:10.4081/ni.2016.6330

3 Badhiwala JH, Ahuja CS, Akbar MA, et al. Degenerative cervical myelopathy — update and future directions. Nat Rev Neurol 2020; 16:108–24. doi:10.1038/s41582-019-0303-0

4 Ito T, Oyanagi K, Takahashi H, et al. Cervical spondylotic myelopathy: Clinicopathologic study on the progression pattern and thin myelinated fibers of the lesions of seven patients examined during complete autopsy. Spine (Phila Pa 1976) 1996; 21:827–33. doi:10.1097/00007632-199604010-00010

5 Seif M, David G, Huber E, et al. Cervical cord neurodegeneration in traumatic and non-traumatic spinal cord injury. J Neurotrauma 2019;:neu.2019.6694. doi:10.1089/neu.2019.6694

6 David G, Seif M, Huber E, et al. In vivo evidence of remote neural degeneration in the lumbar enlargement after cervical injury. Neurology 2019; 92:E1367–77. doi:10.1212/WNL.0000000000007137

7 Grabher P, Mohammadi S, Trachsler A, et al. Voxel-based analysis of grey and white matter degeneration in cervical spondylotic myelopathy. Sci Rep 2016; 6:24636. doi:10.1038/srep24636

8 Martin AR, De Leener B, Cohen-Adad J, et al. Clinically feasible microstructural MRI to quantify cervical spinal cord tissue injury using DTI, MT, and T2∗- weighted imaging: Assessment of normative data and reliability. Am J Neuroradiol 2017; 38:1257–65. doi:10.3174/ajnr.A5163

9 Chen X, Kong C, Feng S, et al. Magnetic resonance diffusion tensor imaging of cervical spinal cord and lumbosacral enlargement in patients with cervical spondylotic myelopathy. J Magn Reson Imaging 2016; 43:1484–91. doi:10.1002/jmri.25109

10 Seif M, Gandini Wheeler-Kingshott CA, Cohen-Adad J, et al. Guidelines for the conduct of clinical trials in spinal cord injury: Neuroimaging biomarkers. Spinal Cord 2019; 57:717–28. doi:10.1038/s41393-019-0309-x

11 Alonso-Ortiz E, Gros C, Foias A, et al. Quantitative MRI of the spinal cord: Reproducibility and normative values across 40 sites. Proc ISMRM SMRT Virtual Conf Exhib 2020.

12 Tetreault L, Kopjar B, Nouri A, et al. The modified Japanese Orthopaedic Association scale: establishing criteria for mild, moderate and severe impairment in patients with degenerative cervical myelopathy. Eur Spine J 2017; 26:78–84. doi:10.1007/s00586-016-4660-8

13 Nurjck S. THE PATHOGENESIS OF THE SPINAL CORD DISORDER ASSOCIATED WITH CERVICAL SPONDYLOSIS. Brain 1972; 95:87–100. doi:10.1093/brain/95.1.87

14 Kirshblum SC, Burns SP, Biering-Sorensen F, et al. International standards for neurological classification of spinal cord injury (Revised 2011). J. Spinal Cord Med. 2011; 34:535–46. doi:10.1179/204577211X13207446293695

15 Curt A, Van Hedel HJA, Klaus D, et al. Recovery from a Spinal Cord Injury: Significance of Compensation, Neural Plasticity, and Repair. J Neurotrauma 2008; 25:677–85. doi:10.1089/neu.2007.0468

16 Yiannakas MC, Kakar P, Hoy LR, et al. The Use of the Lumbosacral Enlargement as an Intrinsic Imaging Biomarker: Feasibility of Grey Matter and White Matter Cross-Sectional Area Measurements Using MRI at 3T. PLoS One 2014; 9:e105544. doi:10.1371/journal.pone.0105544

17 Nouri A, Tetreault L, Zamorano JJ, et al. Role of magnetic resonance imaging in predicting surgical outcome in patients with cervical spondylotic myelopathy. Spine (Phila Pa 1976) 2015; 40:171–8. doi:10.1097/BRS.0000000000000678

18 Fehlings MG, Rao SC, Tator CH, et al. The optimal radiologie method for assessing spinal canal compromise and cord compression in patients with cervical spinal cord injury part II: Results of a multicenter study. Spine (Phila Pa 1976) 1999; 24:605–13. doi:10.1097/00007632-199903150-00023

19 De Leener B, Lévy S, Dupont SM, et al. SCT: Spinal Cord Toolbox, an open-source software for processing spinal cord MRI data. Neuroimage 2017; 145:24–43.https://www.sciencedirect.com/science/article/pii/S1053811916305560 (accessed 25 Oct 2018).

20 De Leener B, Fonov VS, Collins DL, et al. PAM50: Unbiased multimodal template of the brainstem and spinal cord aligned with the ICBM152 space. Neuroimage 2018; 165:170–9. doi:10.1016/j.neuroimage.2017.10.041

21 Ashburner J, Ridgway GR. Symmetric diffeomorphic modeling of longitudinal structural MRI. Front Neurosci 2013; 6:197. doi:10.3389/fnins.2012.00197

22 Mohammadi S, Freund P, Feiweier T, et al. The impact of post-processing on spinal cord diffusion tensor imaging. Neuroimage 2013; 70:377–85. doi:10.1016/j.neuroimage.2012.12.058

23 Mohammadi S, Möller HE, Kugel H, et al. Correcting eddy current and motion effects by affine whole-brain registrations: Evaluation of three-dimensional distortions and comparison with slicewise correction. Magn Reson Med 2010; 64:1047–56. doi:10.1002/mrm.22501

24 David G, Freund P, Mohammadi S. The efficiency of retrospective artifact correction methods in improving the statistical power of between-group differences in spinal cord DTI. Neuroimage 2017; 158. doi:10.1016/j.neuroimage.2017.06.051

25 Tustison NJ, Avants BB. Explicit B-spline regularization in diffeomorphic image registration. Front Neuroinform Published Online First: 2013. doi:10.3389/fninf.2013.00039

26 Sohn SY, Seo JH, Min Y, et al. Changes in dermatomal somatosensory evoked potentials according to stimulation intensity and severity of carpal tunnel syndrome. J Korean Neurosurg Soc 2012; 51:286–91. doi:10.3340/jkns.2012.51.5.286

27 Kramer JK, Taylor P, Steeves JD, et al. Dermatomal somatosensory evoked potentials and electrical perception thresholds during recovery from cervical spinal cord injury. Neurorehabil Neural Repair 2010; 24:309–17. doi:10.1177/1545968309348312

28 Martin A, Leener B De, Cohen-Adad J, et al. Monitoring for Myelopathic Progression with Multiparametric Quantitative MRI. In: Toft M, ed. 2017 CSRS Annual Meeting. Public Library of Science 2017. e0195733. doi:10.1371/journal.pone.0195733

29 Martin AR, Aleksanderek I, Cohen-Adad J, et al. Translating state-of-the-art spinal cord MRI techniques to clinical use: A systematic review of clinical studies utilizing DTI, MT, MWF, MRS, and fMRI. NeuroImage Clin 2016; 10:192–238. doi:10.1016/j.nicl.2015.11.019

30 Uchida K, Baba H, Maezawa Y, et al. Histological investigation of spinal cord lesions in the spinal hyperostotic mouse (twy/twyl: Morphological changes in anterior horn cells and immunoreactivity to neurotropic factors. J Neurol 1998; 245:781–93. doi:10.1007/s004150050287

31 Firooznia H, Ahn JH, Rafii M, et al. Sudden quadriplegia after a minor trauma. The role of preexisting spinal stenosis. Surg Neurol 1985; 23:165–8. doi:10.1016/0090-3019(85)90337-4

32 Wr Y, T L, Tr K, et al. Human neuropathological and animal model evidence supporting a role for Fas-mediated apoptosis and inflammation in cervical spondylotic myelopathy. Brain 2011; 134. doi:10.1093/BRAIN/AWR054

33 Karadimas SK, Moon ES, Yu WR, et al. A novel experimental model of cervical spondylotic myelopathy (CSM) to facilitate translational research. Neurobiol Dis 2013; 54:43–58. doi:10.1016/j.nbd.2013.02.013

34 David G, Mohammadi S, Martin AR, et al. Traumatic and nontraumatic spinal cord injury: pathological insights from neuroimaging. Nat Rev Neurol 2019; 15:718–31. doi:10.1038/s41582-019-0270-5

35 Zhang Z, Guth L. Experimental spinal cord injury: Wallerian degeneration in the dorsal column is followed by revascularization, glial proliferation, and nerve regeneration. Exp Neurol 1997; 147:159–71. doi:10.1006/exnr.1997.6590

36 Mädler B, Drabycz SA, Kolind SH, et al. Is diffusion anisotropy an accurate monitor of myelination?. Correlation of multicomponent T2 relaxation and diffusion tensor anisotropy in human brain. Magn Reson Imaging 2008; 26:874–88. doi:10.1016/j.mri.2008.01.047

37 Buss A, Brook GA, Kakulas B, et al. Gradual loss of myelin and formation of an astrocytic scar during Wallerian degeneration in the human spinal cord. Brain 2004; 127:34–44. doi:10.1093/brain/awh001

38 David G, Weiskopf N, Thompson A, et al. Traumatic and nontraumatic spinal cord injury: pathological insights from neuroimaging. Nat Rev Neurol Published Online First: 2019. doi:10.1038/s41582-019-0270-5

39 Bonizzato M, Pidpruzhnykova G, DiGiovanna J, et al. Brain-controlled modulation of spinal circuits improves recovery from spinal cord injury. Nat Commun2018;9. doi:10.1038/s41467-018-05282-6

40 Courtine G, Sofroniew M V. Spinal cord repair: advances in biology and technology. Nat. Med. 2019; 25:898–908. doi:10.1038/s41591-019-0475-6

